# Prevalence and antibiotic susceptibility patterns of *Staphylococcus aureus* isolated from wounds of diabetic patients attending the Moi Teaching and Referral Hospital

**DOI:** 10.1101/2025.04.07.25325025

**Authors:** D. O. Oguda, S Rono, B. N. Aloo

## Abstract

*Staphylococcus aureus* is a common cause of delayed wound healing worldwide, particularly among diabetic patients, due to the bacterium’s resistance to antibiotics. This study aimed to investigate the prevalence and antimicrobial susceptibility patterns of S. *aureus* isolated from diabetic wound infections at Moi Teaching and Referral Hospital (MTRH). A purposive sampling method was used to select 156 diabetic patients, aged 13 years and above, attending the diabetic clinic at MTRH. Wound swabs were collected aseptically, inoculated onto blood agar, and sub-cultured on Mannitol Salt Agar. The isolates were identified through biochemical tests, and antimicrobial susceptibility was determined using the agar disk diffusion method. Of the 156 samples, 31 (19.87%) were positive for S. *aureus,* while 125 (80.13%) were negative. Among the positive isolates, 26 (10.48%) exhibited intermediate sensitivity, and 72 (29.03%) showed resistance to at least one antibiotic. More than half of the isolates were susceptible to the tested antibiotics. The highest susceptibility was observed for Cefoxitin (96.77%) and Clindamycin (80.65%), while Ampicillin demonstrated the lowest susceptibility (25.81%). The study established, 19.87% prevalence of *S. aureus* in wounds of diabetic patients at the outpatient diabetic clinic of MTRH, with most isolates showing susceptibility to Cefoxitin, Erythromycin, and Clindamycin. Regular surveillance, early screening, and re-evaluation of treatment options, particularly Ampicillin, are essential for effective management diabetic wound infections and to combat antibiotic resistance.

## 1. Introduction

Diabetes is an expensive health issue for both the patient with diabetes and the healthcare systems globally. The number of individuals universally with diabetes has almost multiplied in the previous 40 years (1). According to the International Diabetes Federation (2021), about 463 million of the global population of adults are affected by diabetes, and this figure is expected to increase by at least 1.5 folds by the year 2045 (2). The worldwide incidence of diabetes among adults over 18 years of age has also grown from 4.7% in 1980 to 8.5% in 2014 (3). Diabetes mellitus is a common chronic disease, characterized by persistent hyperglycemia. One of the most serious complications of this disease is diabetic wound infections (4). According to Hurlow (5), diabetic wound infections that contribute substantially to morbidity, prolonged hospital stays, and costs of healthcare globally for diabetes mellitus patients. Approximately up to 25% of diabetics advance into diabetic wound infections in their lifetime. A recent meta-analysis has shown a greater mortality rate in diabetics with diabetic wound infections (99.9 per 1000 person-year) compared to those without diabetic wound infections (41.6 per 1000 person-year)(5).

Diabetic wound infections can contain single or multiple microbes that complicate treatment (6). *Staphylococcus aureus*, are the predominant organism responsible for acute diabetic wound infections (7). *S. aureus* has emerged as a leading causative agent due to its adaptability and multiple virulence factor such as adhesins that facilitate host infection (8), formation of biofilms, and the formation of a polysaccharide capsule and several lytic enzymes that protect it from the host immune system and antibiotics (9–11). This array of virulence factors, therefore, makes the presence of *S. aureus* in diabetic wounds a big challenge in the treatment and management of diabetes. The clinical significance of the pathogen has been exacerbated by the emergence and rapid spread of multidrug resistance among its strains which complicates treatment for people with diabetes (12). According to Anafo (13), the rise of antibiotic-resistant strains such as methicillin-resistant *S. aureus* (MRSA) has especially exacerbated the burden of diabetic wound infections, slowing down their healing rates and commonly resulting in amputations. These resistant strains often lead to treatment failures, necessitating the use of more expensive or toxic antibiotics, thereby increasing the economic and clinical burden on healthcare systems (13).

In Kenya, the presence of *S. aureus* in diabetic wounds has previously been reported by (14–16). The studies have shown that *S. aureus* is a predominant pathogen in diabetic wound infections, contributing to delayed healing and increased complications. Although some reports have indicated emerging antibiotic resistance among these isolates, the scope of existing studies remains limited, with insufficient focus on comprehensive antibiotic susceptibility testing and larger patient populations. Thus, understanding the prevalence and antibiotic susceptibility patterns of *S. aureus* is therefore crucial for tailoring effective treatment strategies and guiding global antibiotic stewardship programs. The present study sought to determine the prevalence and antibiotic susceptibility patterns of *S. aureus* in samples from diabetic wounds from patients attending the Moi Teaching and Referral Hospital (MTRH). The findings of this study can be instrumental in guiding clinical treatment protocols concerning the most effective antibiotics for managing diabetic wound infections. Furthermore, the results can offer critical insights into local antibiotic resistance patterns of *S. aureus*, thereby assisting public health authorities in refining antibiotic stewardship strategies. Ultimately, the results of this study can contribute to improving patient treatment outcomes by reducing the burden of antibiotic-resistant infections, and shaping the overall management of diabetic wounds in Kenya and beyond.

## 2. Materials and methods

### 2.1 Study area and design

The study was conducted at the MTRH. This is a government hospital that is located 310 km northwest of Nairobi in Uasin Gishu County (Eldoret). Several clinics are run at the hospital and the diabetic outpatient clinic is one of these clinics. This was the means of selecting a primary unit for data collection and analysis which was appropriate to specific research questions, hence purposive sampling was used to select patients with diabetic wound infections, the design was appropriate to the study since it helped in gathering baseline information concerning antibiotic susceptibility pattern bacterial diabetic wound infections in diabetic patients at the MTRH. Before the recruitment of participants to the study, their consent was sort through use of Assent forms for those who were between the age of 13-17, while consent forms were used for those who were 18 years and above.

### 2.2 Target population and sample size determination

The target population comprised of T2MD patients who developed diabetic wound infections spinning across 13 years old and above, who visited MTRH diabetic clinic for dressing during the study period from 22^nd^ August 2024 to 31^st^ January 2025. Sample size for the study was determined following Fisher (17) formula as modified by Jung (18) and determined to be 156.

### 2.3 Sample collection and processing

Data on the prevalence and antimicrobial susceptibility of *S. aureus* was collected using a laboratory request form, while socio-demographic data was gathered through a questionnaire administered to participants. Pus specimens from diabetic foot infections were collected by swabbing the wounds aseptically for *S. aureus* screening. The wounds were cleaned with sterile saline, and the swab was moistened with sterile saline before being applied to the wound in a‘zig-zag’ motion to swab the entire surface of the wound. Pus specimens from diabetic foot infections were also collected by swabbing the wounds aseptically for *S. aureus* screening. Gram staining was performed to identify the organisms present in the specimens. The samples were then inoculated onto Blood Agar (BA) plates and incubated at 37°C for 24 to 48 hours. Isolated colonies were sub-cultured onto Mannitol Salt Agar (MSA) and tested for free coagulase enzyme production using the tube coagulase test. All confirmed *S. aureus* strains were further tested for antimicrobial susceptibility using the agar disk diffusion method, following the Clinical and Laboratory Standards Institute (CLSI) 2020 guidelines. Antibiotics tested included; Amoxicillin (30 μg), Ampicillin (10 μg), Cefoxitin (30 μg), Ciprofloxacin (5 μg), Clindamycin (2 μg), Erythromycin (15 μg), Tetracycline (30 μg), and Trimethoprim (25 μg). All experiments were conducted in triplicates to ensure reliability. The results were shared with the participants and the attending clinicians for further management.

### 2.4 Ethical considerations

The authority to conduct research was obtained from NACOSTI (license no: NACOSTI/P/24/34462). Ethical approval to research human subjects was sought from the MTRH/Moi University Institutional Ethics Review Committee (reference no: IREC/895/2024). The purpose of the study was explained to each of the participants. Informed consent was also obtained from patients who meet the desired criteria and who have agreed to participate in the study.

### 2.5 Statistical analysis

Data from the questionnaire and laboratory results was coded and converted all into numerical data which was entered into SPSS version 20. ANOVA was used to determine inferential statistical significance between datasets at 95% confidence level (p ≤ 0.05) and considered to be statistically significant if the difference in antimicrobial performance between the tested isolates and the controls had p values of p ≤ 0.05. Pearson Chi-square (χ2) was used to determine if the risk factors were significantly associated with *S. aureus* infection of wounds from diabetic patients attending outpatient diabetic clinic at MTRH.

## 3. Results

### 3.1 Demographic characteristics of participants

This study utilised A total of 156 patients’ specimens were collected of which 93 (59.62%) were males and 63 (40.38%) were female. Most of the study participants were between the age groups of 45–60 years. A majority (122) of the diabetic patients in this study period were married. 19 (12.18%) were single while 9.62% (15) of the positive cases were from widows/widowers. 69 (44.23%) of the patients had primary school education while 36(23.08%) had secondary school education. 33 (21.15%) of the respondence had tertiary school education with only 18 (11.54%) having no school experience. More than half (56.41%) of the diabetic patients attending outpatient diabetic clinic at the MTRH during the duration of this study had underlying conditions. A majority of the diabetic patients 133 (85.26%) in this study also had previously been hospitalized. A total of 50 (32.05%) had used antibiotics before they enrolled to this study. The data is presented below.

**Table 1.** Demographic characteristics of participants.

| **Factor** | **Categories** | **Total (%)** |
| --- | --- | --- |
| **Age group (Years)** | 13 – 30 | 15 (9.62) |
|  | 31 – 44 | 26 (16.67) |
|  | 45 – 60 | 60 (38.46) |
|  | >60 | 55 (35.25) |
| **Sex** | Female | 63 (40.38) |
|  | Male | 93 (59.62) |
| **Underlying conditions** | Yes | 88 (56.41) |
|  | No | 68 (43.59) |
| **Hospitalized** | Yes | 133 (85.26) |
|  | No | 23 (14.74) |
| **Antibiotics use** | Yes | 50 (32.05) |
|  | No | 106 (67.95) |
| **Marital Status** | Single | 19 (12.18) |
|  | Married | 122 (78.21) |
|  | Other | 15 (9.62) |
| **Level of Education** | Primary | 69 (44.23) |
|  | Secondary | 36 (23.08) |
|  | Tertiary | 33 (21.15) |
|  | No School | 18 (11.54) |

### 3.2 Prevalence of *Staphylococcus aureus* from diabetic wounds of patients

This study utilised 156 (100%) samples obtained from wounds of diabetic patients attending outpatient diabetic clinic at the MTRH during the period from August 2024 to January 2025. The swabs collected were aseptically inoculated into Tryptone Soy broth and thereafter sub-cultured on blood agar. Brown or white, beta-hemolytic, and round colonies obtained were then sub-cultured on Mannitol Salt Agar (MSA) for identification (Fig 1). The yellowish appearance of colonies on MSA indicated presence of *S. aureu*s.

**Fig 1.**
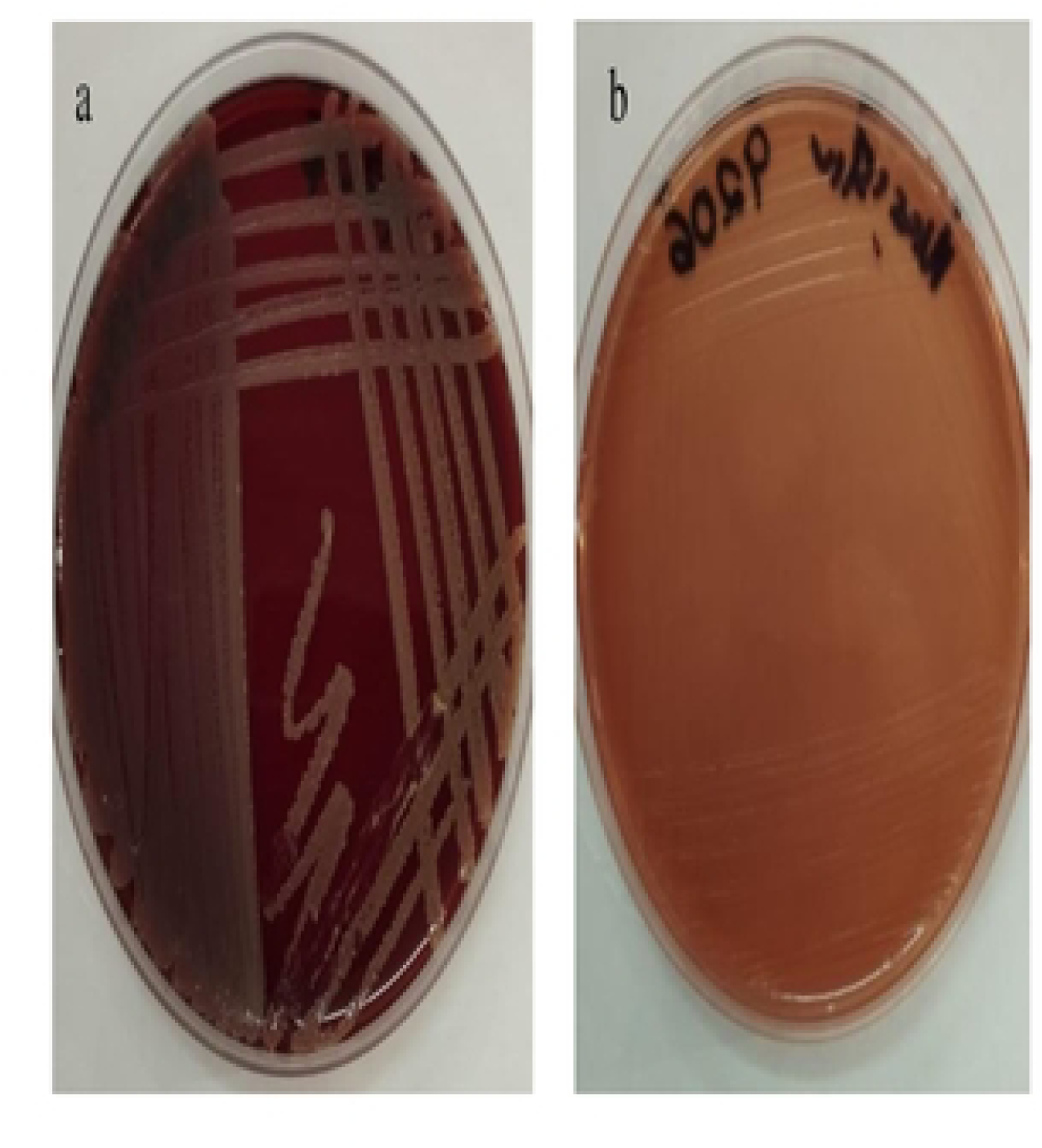
*Staphylococcus* species on Chocolate agar (a) and Mannitol Salt agar (b)

A loopful of each distinct colony was subjected to gram staining and morphological identification revealed by microscopy. Purple, cocci-shaped cells arranged in pairs and clusters were identified as *S. aureus* (Fig 2a). Further identification was obtained through biochemical testing using Catalase (Fig 2b) and Coagulase (Fig 2c).

**Fig 2.**
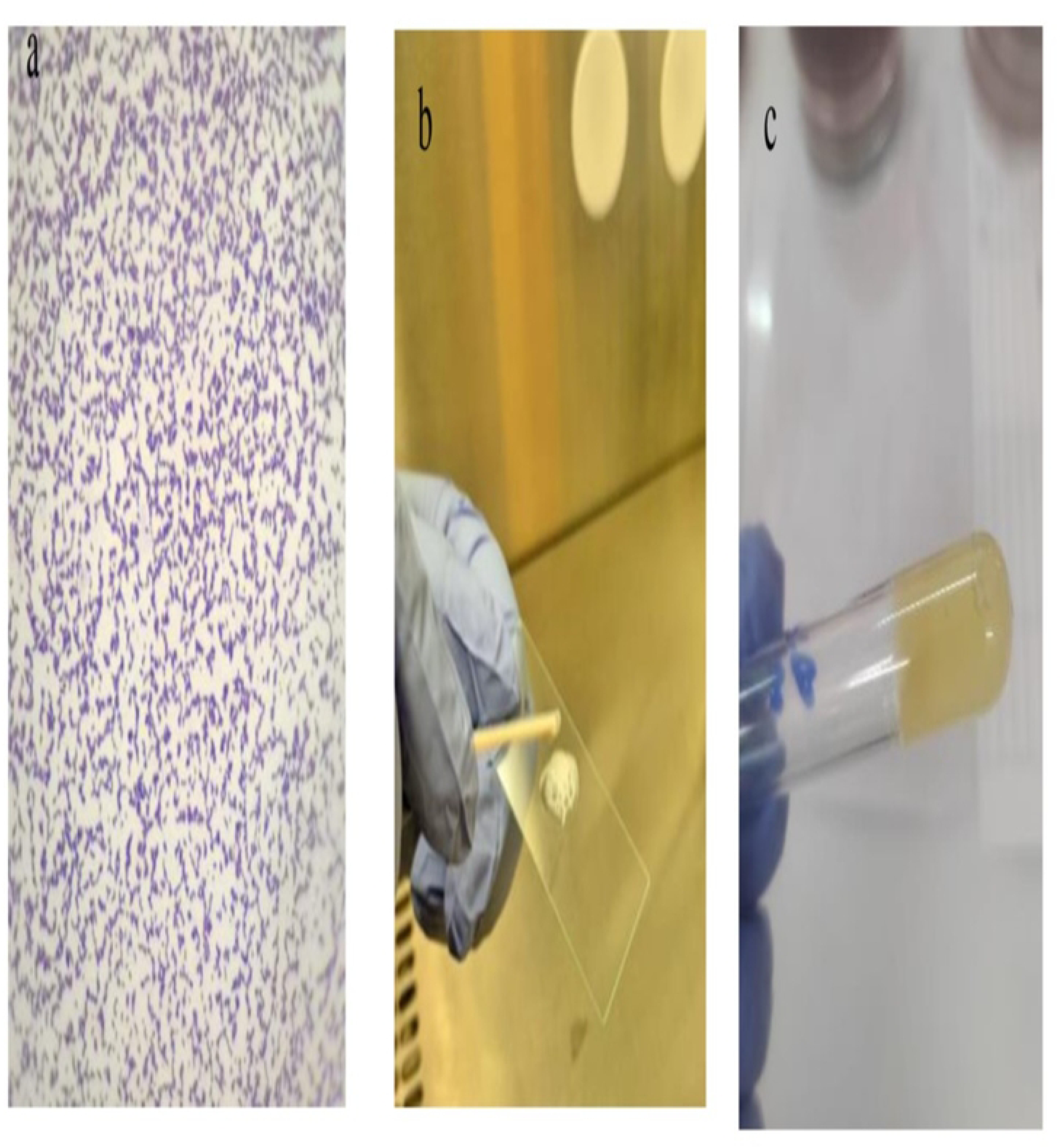
Further identification of *S. aureus* by Gram Staining (a), Catalase test (b) and Coagulase test (c)

Upon confirmation of *S. aureus* from the samples, this study noted that 31 (19.87%) samples were positive while 125(80.13%) samples tested negative. This translates to a prevalence of 19.87% as shown in Table 2.

**Table 2:**
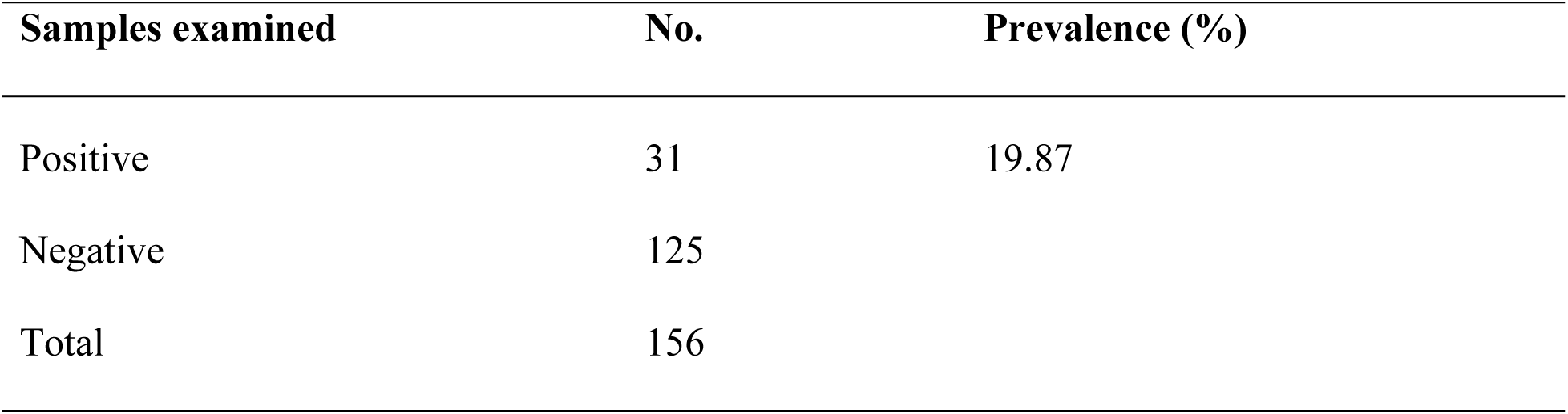
Prevalence of *Staphylococcus aureus* from wounds of diabetic patients at MTRH Samples examined.

| Samples examined | No. | Prevalence (%) |
| --- | --- | --- |
| Positive | 31 | 19.87 |
| Negative | 125 |  |
| Total | 156 |  |

### 3.3 Antibiotic susceptibility patterns of the *S. aureus* isolates

This study assessed the antibiotic susceptibility patterns of the isolated *S. aureus* against eight (8) antibiotics of different classes/families and with different modes of action as prioritized under the MTRH protocol. The tested antibiotics were Amoxicillin (30 μg), Ampicillin (10μg), Cefoxitin (30μg), Ciprofloxacin (5μg), Clindamycin (2μg), Erythromycin (15μg), Tetracycline (30μg) and Trimethoprim (25μg). The clear zones formed around the discs were recorded as zones of inhibition (Fig 3) which were measured in millimetres using vernier callipers. *S. aureus* had varying degrees of susceptibility profiles to the antibiotics they were subjected to by disc diffusion method (Table 3 & Fig 4). *Staphylococcus aureus* isolated in this study isolated bacteria had at least one instance of intermediate sensitivity 26 (10.48%) and/or antibiotic resistance 72 (29.03%) to the other antibiotics. However, more than half of the isolates were susceptible to the test antibiotics as shown in Table 3. Higher number of *S. aureus* isolates were susceptible to Cefoxitin (96.77%) and Clindamycin (80.65%) with lesser susceptibility to Ampicillin (25.81%) (Table 3).

**Fig 3.**
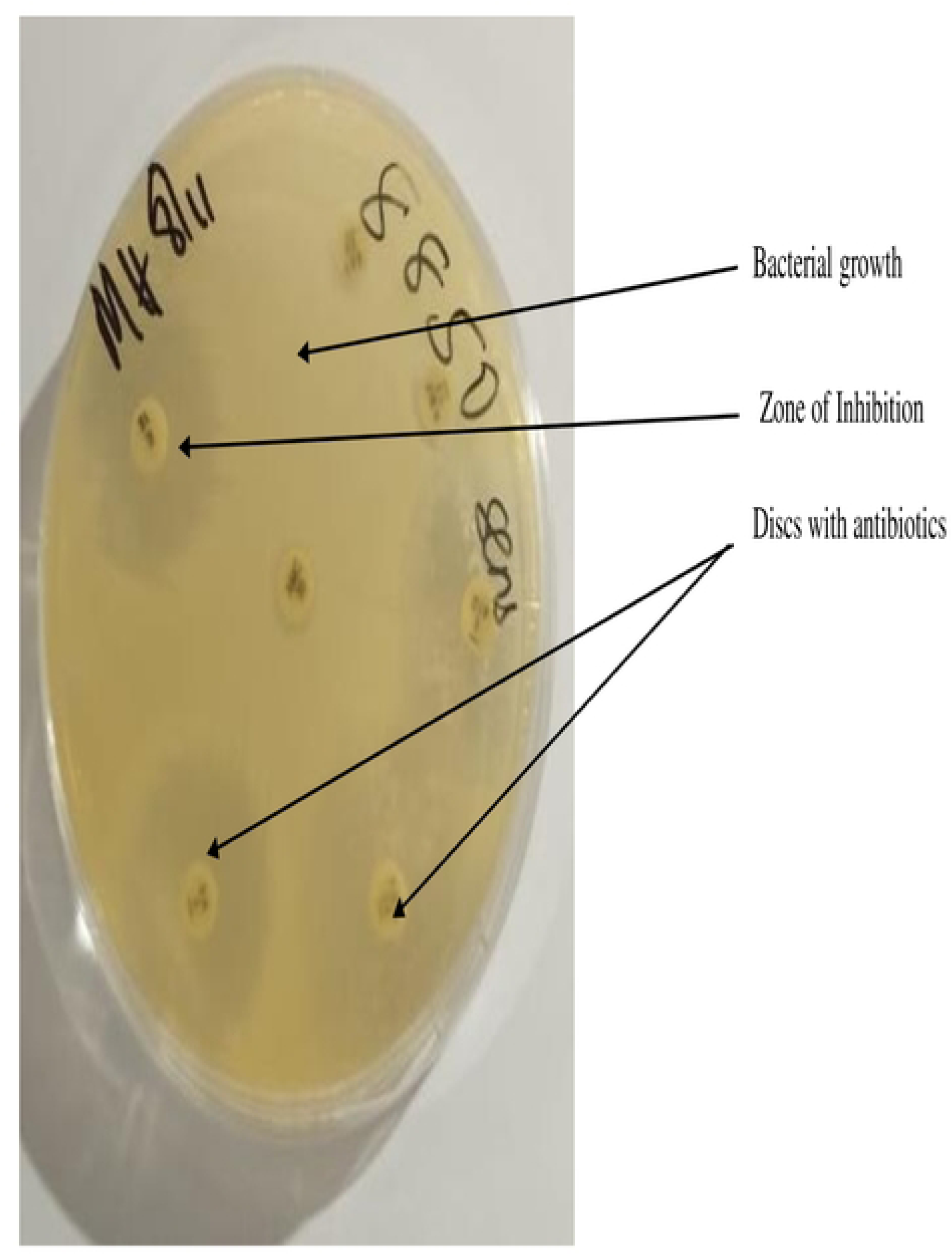
Antibacterial susceptibility test of *S. aureus* showing clear zones of inhibition.

**Fig 4.**
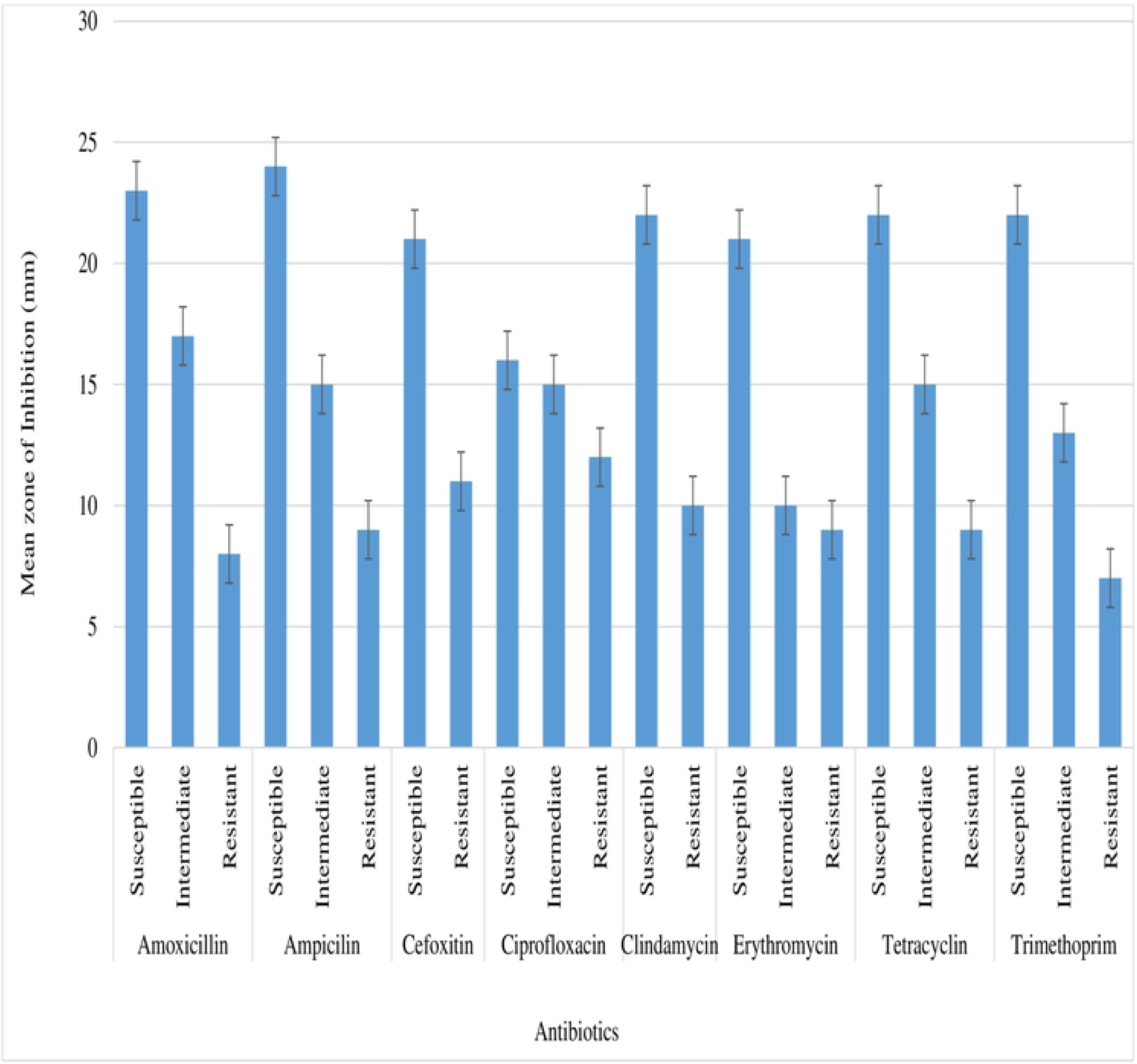
Antibacterial susceptibility profiles of *Staphylococcus aureus* isolates from wounds of diabetic patients attending outpatient diabetic clinic at MTRH.

**Table 3:** Susceptibility profile of *S. aureus* to tested antibiotics.

| Antibiotic | Susceptible N (%) | Intermediate N (%) | Resistant N (%) |
| --- | --- | --- | --- |
| Amoxicillin | 18 (58.06) | 5 (16.13) | 8 (25.81) |
| Ampicillin | 8 (25.81) | 11 (35.48) | 12 (38.71) |
| Cefoxitin | 30 (96.77) | - | 1 (3.23) |
| Ciprofloxacin | 16 (51.61) | 2 (6.45) | 13 (41.94) |
| Clindamycin | 25 (80.65) | - | 6 (19.35) |
| Erythromycin | 20 (64.51) | 1 (3.23) | 10 (32.26) |
| Tetracycline | 15 (48.39) | 6 (19.35) | 10 (32.26) |
| Trimethoprim | 18 (58.06) | 1 (3.23) | 12 (38.71) |

ANOVA was used determine the susceptibility of *S. aureus* isolated from diabetic wounds of patients attending outpatient diabetic clinics at the MTRH. The statistically significant p-value of 0.0000 at 95% confidence level (P ≤ 0.05) (Table 4) means that the *S. aureus* isolates are statistically susceptible to test antibiotics.

**Table 4:**
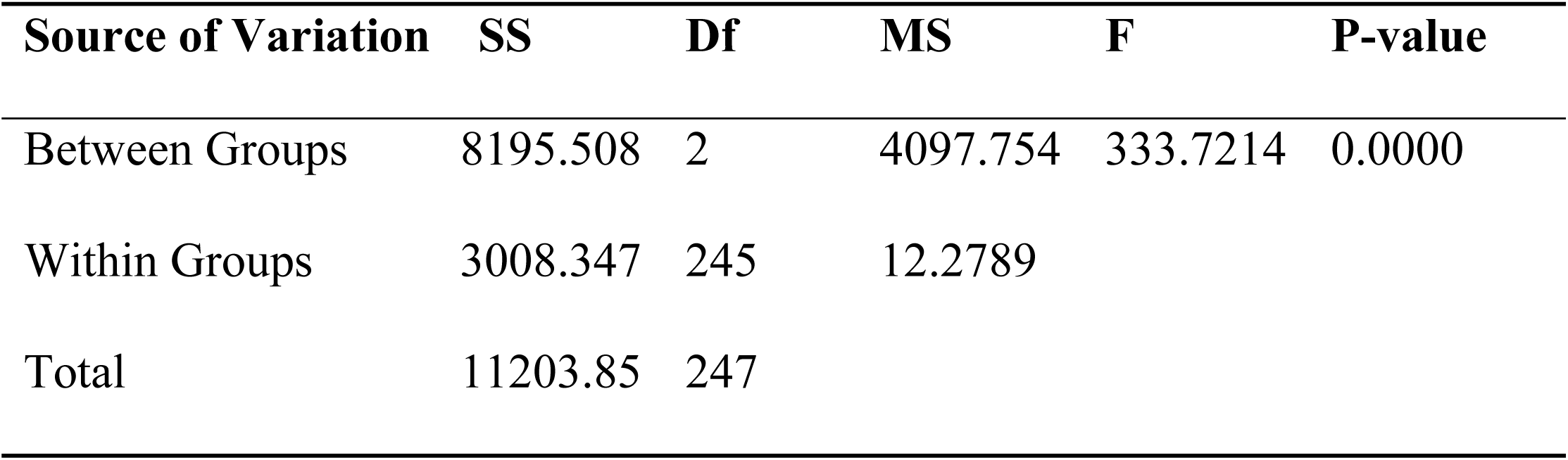
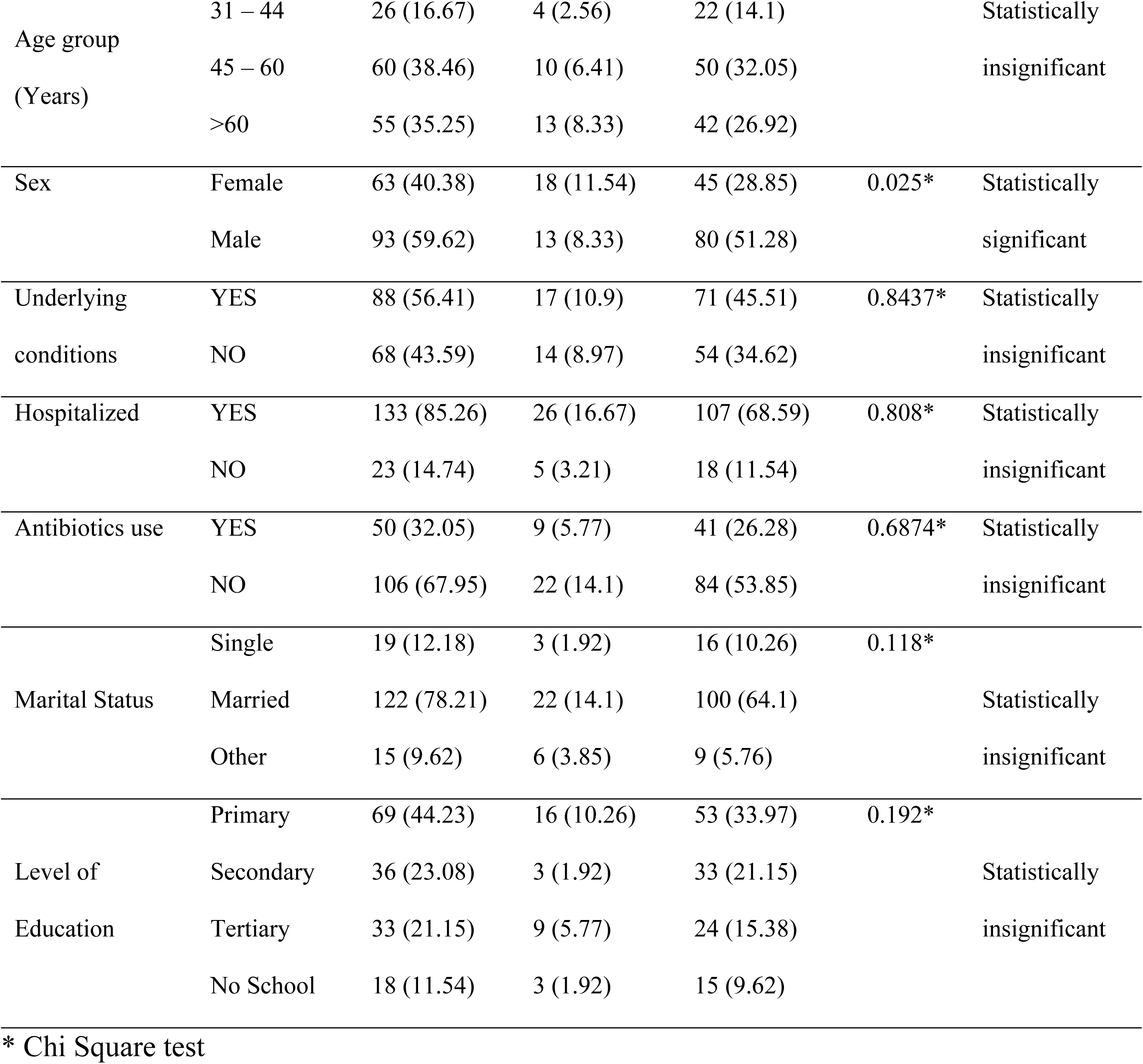
ANOVA table on susceptibility of *S. aureus* isolated from diabetic wounds of patients attending outpatient diabetic clinics at MTRH.

### 3.3 Risk factors associated with *S. aureus* infection of wounds from diabetic patients attending outpatient diabetic clinic at MTRH

Out of the 31 (19.87%) positive cases for presence of *S. aureus* in wounds of diabetic patients, majority (13) were patients more than 60 years of age. Age groups of 13 – 30 and 31 – 44 had 4(2.56%) positive cases each, while those aged between 45 and 60 years old were 10(6.41%) (Table 4). Despite those aged more than 60 years returning a higher positivity rate, the statistically insignificant Chi-square test p-value of 0.6503 at 95% confidence level indicated that the age of a diabetic patient(s) is not directly linked to presence of *S. aureus* in their wounds (Table 5). This means that *S. aureus* is likely to be present in wounds of diabetic patients attending outpatient diabetic clinic at MTRH irrespective of their age.

**Table 5.**
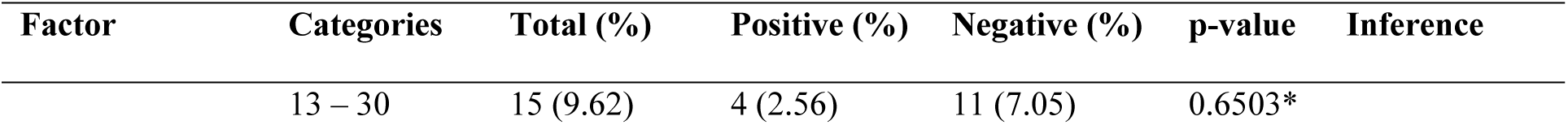
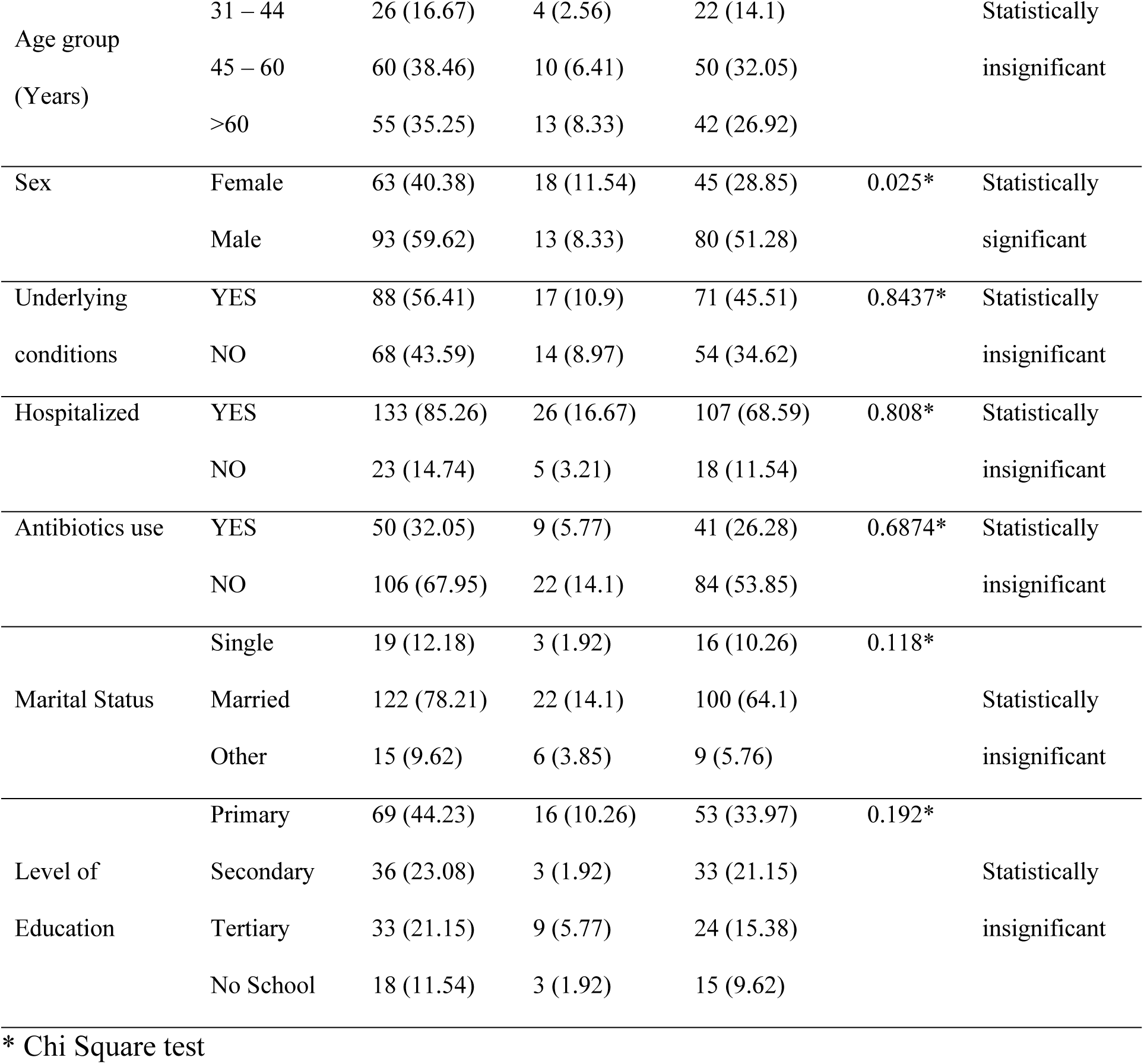
Risk factors associated with *S. aureus* infection of wounds from diabetic patients at the MTRH.

A total of 93 (59.62%) male and 63 (40.38%) female diabetic patients attending outpatient diabetic clinic at MTRH were enrolled in this study. Of the 31 positive cases, 13 (8.33%) were male while 18 (11.54%) were female. The statistically significant chi-square test p-value of 0.025 at P ≤ 0.05 means that the sex of a diabetic patient significantly influences infection with *S. aureus* (Table 5). The sex of a diabetic patients particularly those attending outpatient diabetic clinic at the MTRH can be linked to existence and subsequent isolation of *S. aureus* in their wounds. This is evident from the results of this study whereby female diabetic patients returned a significantly higher positivity rate when compared to male diabetic patients.

More than half (56.41%) of the diabetic patients attending outpatient diabetic clinic at MTRH during the duration of this study had underlying conditions. Out of the total (31) positive cases of *S. aureus* infection, 17 (10.9%) were from diabetic patients who had underlying conditions with those without accounting for only 8.97% (14) of the total patients enrolled (Table 5). Using chi-square test, the statistically insignificant p-value of 0.8437 at 95% confidence level indicates that there was no relation between existence of underlying conditions and presence of *S. aureus* in wounds (Table 5). That means that diabetic patients are likely to get *S. aureus* infection whether they have underlying conditions or not.

Majority of the diabetic patients 133 (85.26%) in this study had previously been hospitalized for various reasons out of which 26 (16.67%) of them returned a positive result (Table 5). Despite that, the statistically insignificant chi-square test p-value of 0.808 at 95% confidence level indicates that there was no relation between existence of previous hospitalization and presence of *S. aureus* in wounds (Table 5). This infers those diabetic patients attending outpatient diabetic clinic at MTRH are likely to have *S. aureus* in their wounds irrespective of their prior hospitalization status.

A total of 50 (32.05%) diabetic patients attending outpatient diabetic clinic at MTRH in this study had used antibiotics prior to enrolment. Of the 31 (19.87%) positive cases, 9 (5.77%) diabetic patients had used antibiotics while 41 (26.28%) had no history preceding their enrolment (Table 5). The statistically insignificant chi-square test p-value of 0.6874 at 95% confidence level indicates that there was no relation between prior antibiotic use and the presence of *S. aureus* in the wounds of diabetic patients (Table 5).

A majority (122) of the diabetic patients attending outpatient diabetic clinic at the MTRH during this study period were married. 14.1% (22) of them returned positive results for *S. aureus* infections in their wounds. Three (1.92%) of the positive cases were from those who were single while 3.85% (6) of the positive cases were from others (widows/widowers) (Table 5). Despite more married returning a higher positivity rate, the statistically insignificant Chi-square test p-value of 0.118 at 95% confidence level indicates that marital status of a diabetic patient(s) is not directly linked to presence of *S. aureus* in their wounds (Table 5). This means that *S. aureus* is likely to be present in wounds of diabetic patients at MTRH irrespective of their marital status.

Out of the 31 (19.87%) positive cases for presence of *S. aureus* in wounds of diabetic patients, majority 16 (10.26%) out of 69 had primary school education. Those with tertiary school education were 33 with 9 (5.77%) returning positive results of *S. aureus* infections. Diabetic patients who attended outpatient diabetic clinic at the MTRH during the duration of this study and had no school experience or had with secondary school education returned 3 (1.92%) positive cases each, from a total of 18 and 36 respectively (Table 5). Despite those who had primary school education only having a higher positivity rate, the statistically insignificant Chi-square test p-value of 0.192 at 95% confidence level indicated that the level of education of a diabetic patient(s) is not directly linked to presence of *S. aureus* in their wounds (Table 5). This means that *S. aureus* is likely to be present in wounds of diabetic patients at the MTRH irrespective of their level of education.

## 4. Discussion

A total of 156 samples were obtained from wounds of diabetic patients attending outpatient diabetic clinic (Chandaria Cancer and Chronic Diseases Centre) at the MTRH during the study period. 31 samples were positive while 125 samples tested negative. This translates to a prevalence of 19.87%. The prevalence result of this study is lower when compared to similar studies at conducted at Vihiga County Referral Hospital (14). That study established an overall 60.3 % prevalence of *S. aureus* infection among diabetes mellitus patients. Similar studies in Ethiopia also recorded higher prevalence of 31.1% (19) and 25.19% (20). Mutonga (15) recorded 98% prevalence while Amini (21) reported 87% prevalence. The difference can not only be attributed to the differences in time periods and settings in which the studies were conducted but also the fact that these studies focused on foot ulcers. According to Mutonga (15), it is estimated that 10–15% of diabetic patients will develop DFUs at some point in their life. TUVEI (14) also documents that foot ulcers are more prone to infections than other wounds. These high prevalences can be attributed to the fact that most often, *S. aureus* colonizes on skin or mucosal surfaces. However, it has been documented that children, HIV or diabetic patients are more prone to *S. aureus* colonization if they have wounds (22). They also have potential to cause serious infections if not treated early. Even when the virulence and invasive capability of *S. aureus* strains recovered from diabetes patients’ wounds are lower than those of strains typically seen in infections, they nevertheless retain the ability to cause and sustain invasive and deep tissue infections (23).

*Staphylococcus aureus* isolated in this study had at least one instance of intermediate sensitivity 26 (10.48%) and/or antibiotic resistance 72 (29.03%) to the other antibiotics. However, more than half of the isolates were susceptible to the test antibiotics. This indicates that antibiotic resistance is not widespread among diabetic patients, which has also been reported by other authors before (14,15,19–21).

A higher number of *S. aureus* isolates were susceptible to Cefoxitin, Erythromycin and Clindamycin with lesser susceptibility to Ampicillin. These findings differ from those of Amini (21) reported 63.9% resistance by *S. aureus* isolates to Clindamycin. Atlaw (20) also documented a high level of resistance of *S. aureus* to erythromycin and trimethoprim unlike in the current study. The authors however reported that the *S. aureus* isolates were sensitive to clindamycin just like in the current study. These also concur with the findings of Fawad (24) who documented that *S. aureus* they isolated showed high sensitivity to Cefoxitin and Clindamycin. Bhat (25) also documented that *S. aureus* presented better susceptibility to commonly used antibiotics like Erythromycin, ceftriaxone and clindamycin. *S. aureus* isolates showed high rates of resistance to oxacillin (95.2%), clindamycin (68.7%), erythromycin (65.6%) according to Owais (26). The spread of antibiotic-resistant *S. aureus* complicates therapy, highlighting the necessity of implementing strong infection control methods like improved hygiene and creating novel therapeutic approaches. Given the variability in antibiotic resistance patterns, personalized treatment plans based on susceptibility testing are crucial for effective management.

Out of the 31 (19.87%) positive cases, a majority (8.33%) were patients more than 60 years of age. Age groups of 13 – 30 and 31 – 44 had 4 (2.56%) positive cases each, while those aged between 45 and 60 years old were 10 (6.41%). This concurs with earlier studies as well. TUVEI (14) reported an even higher prevalence rate among those in the age group of over 60 years at 63.8%. Amini (21) also documented that more than half (51/90) were more than 60 years. This was similar to Rashid (27) who established a higher prevalence for those aged over 50 years. This observation could be attributed to these group of patients having other pre-existing medical conditions like hypertension, reduced mobility and rare visits to the diabetic clinic. The danger of T2D is at its peak with an increase in age, especially after 45 years, due to less exercise thus gaining weight (28). Therefore, aging may augment T2DM risk through pathophysiological mechanisms independent of obesity. As has been reported by other authors before, as people age, their immune systems become less effective at fighting off infections. Older individuals often have reduced neutrophil function and other immune impairments, making them more susceptible to infections like *S. aureus* (29).

Of the 31 positive cases in this study, 13 (8.33%) were male while 18 (11.54%) were female. The statistically significant chi-square test p-value of 0.025 means that the sex of a diabetic patient significantly influences infection with *S. aureus*. The sex of a diabetic patients particularly those attending outpatient diabetic clinic at MTRH can be linked to existence and subsequent isolation of *S. aureus* in their wounds. This is evident from the results of this study whereby female diabetic patients returned a significantly higher positivity rate (11.54%) when compared to male diabetic patients. The highest prevalence of *S. aureus* infections in diabetic wounds in females could be attributed to the kind of chores traditionally female dominated. Most studies have not explored the influence of sex in occurrence of diabetes. That is despite an estimated 17.7 million more men than women worldwide suffering from diabetes mellitus. However, when type 2 diabetes is diagnosed, women seem to have a higher load of risk factors. Most studies have also not explored the influence of sex in occurrence of diabetes. Women experience greater hormonal fluctuations throughout their lives, particularly during pregnancy and menopause, which can affect glucose metabolism and increase the risk of developing diabetes (30). The influence of sex is therefore inconclusive as some studies demonstrated male gender as a risk factor, some female gender as a risk factor while other studies have shown no difference. Amini (21) reported equal proportions of infections in both sexes. TUVEI (14) reported that females had a higher prevalence of 57.4% as compared to their male counterparts at 42.6% just as was in this study also were of the same view (27,31,32). However, this study results contradicted those by other studies reporting high prevalence in males than females (20,33–36).

More than half (56.41%) of the diabetic patients attending outpatient diabetic clinic at MTRH during the duration of this study had underlying conditions. Out of the total (31) positive cases of *S. aureus* infection, 17 (10.9%) were from diabetic patients who had underlying conditions. The most frequent was hypertension followed heart disease and kidney disease. Just like other studies before, this study found out that underlying conditions raise diabetes risk by altering metabolic health, causing insulin resistance, and complicating care of modifiable risk factors. The higher positivity rate can be attributed to the already low immune systems among most diabetic patients. Underlying conditions weaken the patient’s immune system rendering them highly susceptible to other infections. Reveles (34) is of the same opinion, with their findings comparable to those from this study. According to the authors hypertension (76%), dyslipidemia (52%), obesity (49%), peripheral vascular disease (37%), and kidney disease (12%) significantly predisposes diabetic patients to *S. aureus* infection.

A majority of the diabetic patients 133 (85.26%) in this study had previously been hospitalized for various reasons out of which 26 (16.67%) of them returned a positive result. This is slightly lower that from the findings of Reveles (34). The authors documented a prevalence of 19% from patients with a history of recent hospitalization. This suggests that it is possible that *S. aureus* infection among diabetic patients in this and other studies could be due to nosocomial risk. The use of improperly sterilized equipment as well as contaminated fomites in the hospitals could be the main reason.

The present study identified previous hospitalization as an independent risk factor for *S. aureus* infection. Hospital-acquired infection is one of the most common causes of most microbial infections (37). According to Liu (38), the occurrence of hospital acquired infection is mainly due to the poor ward environment and the inadequate implementation of isolation measures for patients.

Another 50 (32.05%) diabetic patients attending outpatient diabetic clinic at MTRH in this study had used antibiotics prior to enrolment. Of the 31 (19.87%) positive cases, 9 (5.77%) diabetic patients had used antibiotics. The statistically insignificant chi-square test p-value of 0.6874 at 95% confidence level indicates that there was no relation between prior antibiotic use and the presence of *S. aureus* in the wounds of diabetic patients. This means that the *S. aureus* isolates could not be directly labelled as resistant to the antibiotics used by the patients prior to this study. That means they may have been contracted after completion of the prescribed dosage. However, that contradicts the findings from Amini (21) who reported that 55.4% of the positive cases had a history of recent antibiotic therapy in last few days. Reveles (34) also hold the same view with 43% positivity rate documented by the authors. According to Yuan (39), antibiotics can raise the risk of diabetes by alter the gut microbiota and impact metabolic health. Additionally, the risk may be confounded by underlying diseases that need the use of antibiotics.

A majority (122) of the diabetic patients attending outpatient diabetic clinic at MTRH during this study period were married. 14.1% (22) of them returned positive results for *S. aureus* infections in their wounds. 1.92% (3) of the positive cases were from those who were single while 3.85% (6) of the positive cases were from others (widows/widowers). Similar findings have also been documented by earlier studies. TUVEI (14) noted that those married had a higher prevalence at 84.0%. Aedh (33) also documented similar findings. According to Karimi (40), divorced people are less likely to have T2DM, widowed people are less likely to have the T2DM, and single people are more likely to have it. It has also been documented that single men may have a higher risk of diabetes compared to married men, while the impact on women can be different depending on the specific marital status. Social support, which is frequently provided by marriage, has a favourable impact on health-related behaviours including diet, exercise, and treatment compliance. Regarding the effect of marital status on *S. aureus* infection rates, the majority of studies have produced conflicting findings, with the majority suggesting that married participants had higher rates (41). This can be attributed to increased exposure and transmission opportunities within households.

Out of the 31 (19.87%) positive cases for presence of *S. aureus* in wounds of diabetic patients, majority 16 (10.26%) out of 69 had primary school education. Those with tertiary school education were 33 with 9 (5.77%) returning positive results of *S. aureus* infections. Diabetic patients who attended outpatient diabetic clinic at MTRH during the duration of this study and had no school experience or had with secondary school education returned 3 (1.92%) positive cases each, from a total of 18 and 36 respectively. (14) conducted an education level analysis from their data and noted that those with primary level education had the highest prevalence rate at 51.1% just like in the current study. Aedh (33) also reported similar findings. Reduced socioeconomic status is frequently associated with lower education levels, which can result in more exposure to cramped living arrangements, unsanitary environments, and restricted access to medical treatment. Low level of education may also attribute to poor dressing of their wounds as most do not acquire sanitary techniques. Highly educated people may have easier access to early detection and treatment, which lowers the risk of severe infections (42). These factors can increase the risk of diabetes and/or *S. aureus* infections.

## 5. Conclusions

The current study reports an *S. aureus* prevalence of 19.87% from wounds of diabetic patients attending outpatient diabetic clinic (Chandaria Cancer and Chronic Diseases Centre) at Moi Teaching and Referral Hospital. The sample size was 156 patients with 31 testing positive while 125 tested negatives. The current study concludes that majority of the *S. aureus* isolates in this study were susceptible to Cefoxitin, Erythromycin and Clindamycin with lesser susceptibility to Ampicillin. The other antibiotics had at least one instance of intermediate sensitivity and/or antibiotic resistance by the isolates. The study recommends regular examination and early screening of the most common pathogens in wounds of diabetic patients to get first-hand knowledge about the identification to detect infections early so that healthcare providers can initiate preventive measures. Continued surveillance and periodical monitoring to determine the susceptibility profile of *S. aureus* and other pathogens in wounds of diabetic patients attending outpatient diabetic clinic at the MTRH is recommended. Re-evaluation of treatment options particularly the use of Ampicillin should also be taken into consideration to prevent widespread antibiotic resistance.

## Data Availability

All data produced in the present work are contained in the manuscript

## Acknowledgments

The authors would like to acknowledge Moi Teaching and Referral Hospital diabetic clinic and Laboratory Directorate staff and the entire management for the opportunity to carry out our research within the facility and their kind cooperation and encouragements

## Ethical Considerations

The study was approved before commencement by obtained from NACOSTI (license no: NACOSTI/P/24/34462) as well as the Institutional Research and Ethics Committee (IREC) of Moi MTRH and Moi University School of Medicine and permission to conduct the study was sought from MTRH management (Approval No: **0004852**). A written informed consent was obtained from all participants prior to commencement of the study. Confidentiality was maintained throughout the study using a password protected database and limiting the access only to the principal investigators and the research assistants.

## Data availability

The study was part of a thesis submitted to University of Eldoret. Data will soon be available at the university repository but can also be availed in excel by the corresponding author if need be.

